# ML-Guided GWAS Reveals Genetic Architectures for MASLD for Overweight and Lean Individuals in the *All of Us* Cohort

**DOI:** 10.64898/2025.12.18.25342567

**Authors:** Ananthan Nambiar, Kaushik Karambelkar, Arjun Athreya, Alina M. Allen, Konstantinos N. Lazaridis, Sharon M. Donovan, Sergei Maslov

**Affiliations:** Department of Bioengineering University of Illinois Urbana-Champaign Urbana IL 61801 U.S.A.; Carl R. Woese Institute for Genomic Biology Urbana IL 61801 U.S.A; Center for Individualized Medicine Mayo Clinic Rochester MN 55905 U.S.A.; Department of Genome Sciences University of Washington Seattle WA 98195 U.S.A.; eScience Institute University of Washington Seattle WA 98195 U.S.A.; Department of Molecular Pharmacology and Experimental Therapeutics Mayo Clinic Rochester MN 55905; Department of Psychiatry and Psychology Mayo Clinic Rochester MN 55905; AMA Department of Medicine Division of Gastroenterology and Hepatology Mayo Clinic Rochester MN 55905; Department of Food Science and Human Nutrition University of Illinois Urbana-Champaign Urbana IL 61801 U.S.A.; Personalized Nutrition Initiative University of Illinois Urbana-Champaign Urbana IL 61801 U.S.A.; Department of Physics University of Illinois Urbana-Champaign Urbana IL 61801 U.S.A.; Computing Environment and Life Sciences Argonne National Laboratory Lemont IL 60439 U.S.A.

**Keywords:** Machine Learning, Genome-Wide Association Study, Metabolic Dysfunction-Associated Steatotic Liver Disease, Liver

## Abstract

Metabolic dysfunction-associated steatotic liver disease (MASLD) arises from excessive hepatic fat accumulation that triggers inflammation and liver injury. It is the most prevalent chronic liver disease worldwide, affecting more than one quarter of adults. Despite this, MASLD is often underdiagnosed, making it more difficult to perform genome-wide association studies (GWAS). In this paper, we implemented a machine learning (ML)-guided GWAS framework to identify genetic risk factors for MASLD across lean and overweight individuals in the *All of Us* Research Program. A random forest model trained on laboratory measurements, vital signs, and demographic features generated an in silico MASLD (I-MASLD) score, a continuous risk score for MASLD, which was validated to accurately represent clinical MASLD diagnosis. This score was then used as the phenotype in a GWAS of whole-exome sequencing variants. The resultant GWAS discovered a novel variant in the ANGPTL4 gene to be significantly associated with MASLD risk and recapitulated known variants in various genes involved in lipid metabolism and insulin signaling. Our results also suggest a potential role of *APOA5* in MASLD onset or progression in lean patients. These findings demonstrate that ML-derived quantitative phenotypes can enhance genetic discovery in large, heterogeneous cohorts where conventional case/control labels are limited or imprecise.

## 1. Introduction

Metabolic dysfunction-associated steatotic liver disease (MASLD), previously known as non-alcoholic fatty liver disease (NAFLD), is the most common form of chronic liver disease globally, affecting over a quarter of the adult population (1; 2). Despite its clinical relevance and its association with other diseases including cardiovascular disease and type 2 diabetes, MASLD remains markedly underdiagnosed due to its asymptomatic presentation in early stages and the limited use of liver imaging or biopsy in routine care (3).

The prevalence of MASLD is higher among individuals with obesity, where insulin resistance and dyslipidemia frequently co-occur (4). Although not as prevalent, MASLD can also occur in normal-weight individuals and is commonly referred to as "lean MASLD". Lean patients account for about 10-15% of MASLD-affected individuals (5). Lean MASLD underscores that factors beyond body weight, such as genetic predisposition, metabolic alterations, and lifestyle influences, contribute to hepatic steatosis and disease susceptibility. Despite being extensively investigated in individuals with obesity, the underlying pathogenic mechanisms of lean MASLD remain poorly understood.

MASLD arises from the interplay of metabolic, environmental, and genetic factors that influence hepatic fat accumulation and inflammation. In recent years, genetic studies have revealed that inherited variants can substantially modulate individual susceptibility to MASLD by altering lipid metabolism and hepatic energy homeostasis. For instance, the patatin-like phospholipase domain-containing 3 (*PNPLA3*) gene encodes a triglyceride lipase responsible for mobilizing polyunsaturated fatty acids (PUFAs) for hepatic secretion of large very low-density lipoproteins (VLDL). Its I148M variant—one of the most robust and consistently replicated genetic risk factors for MASLD—impairs this process, resulting in hepatic lipid accumulation (6). In individuals with obesity, these genetic effects may interact with systemic metabolic stress and dietary exposures to further elevate MASLD risk, whereas in individuals without obesity, they may contribute to hepatic steatosis even in the absence of overt metabolic dysfunction. Together, these findings support a clinical scenario in which multiple inherited variants, acting through diverse physiological mechanisms, contribute to MASLD susceptibility in concert with environmental and metabolic stressors.

Recent biobank-scale resources such as the UK Biobank (7) and the *All of Us* Research Program (8; 9) have created unprecedented opportunities to investigate the genetic architecture of MASLD. In the UK Biobank, magnetic resonance imaging–based proton density fat fraction (MRI-PDFF) data has enabled the use of hepatic fat content as a direct, quantitative phenotype for genome-wide association studies (GWAS), leading to the discovery and validation of several MASLD-associated loci including *PNPLA3* and the transmembrane 6 superfamily member 2 (*TM6SF2*) gene (10). In contrast, the *All of Us* cohort lacks imaging- or biopsy-based phenotypes, instead linking whole-genome sequencing with electronic health records (EHRs), self-reported data, and laboratory measurements across a genetically diverse population of Americans. While *All of Us* provides a valuable platform for discovering genetic risk factors in historically underrepresented populations, the absence of deep liver phenotyping presents a major challenge for MASLD GWAS. Specifically, defining MASLD as a binary outcome based on International Classification of Diseases (ICD) codes (e.g., K76.0) does not capture subclinical or undiagnosed cases, a critical limitation given that MASLD is often asymptomatic and rarely detected early in the absence of routine liver screening.

Machine learning (ML) models trained on laboratory measurements, clinical signs, medication history, and demographic features are increasingly employed to predict disease onset across a range of complex conditions. Notably, ML-based risk models have been applied to MASLD, diabetes, chronic kidney disease, and coronary artery disease (11). In contexts where disease status is underdiagnosed or intrinsically continuous, as is the case with MASLD, ML models provide a principled approach to infer latent phenotypes from observable clinical features. When used as phenotypes in GWAS, these ML-derived risk scores directly address the limitations of binary disease labels by providing a continuous measure of disease likelihood, capturing gradations of risk that are often missed in case/control definitions. Recent studies in coronary artery disease and glaucoma have demonstrated that ML-derived phenotypes can enhance statistical power for genetic discovery and facilitate the identification of novel risk loci (12; 13).

In this study, we applied an ML-guided GWAS framework to investigate the genetic architectures of MASLD across individuals with different body compositions using data from the *All of Us* Research Program. We conducted separate genome wide association analyses to identify risk alleles specifically associated with over-weight (BMI > 25) and lean (BMI < 25) MASLD to identify alleles that confer risk within each group. First, we trained an ML model on non-genomic clinical features, including laboratory values, vital signs, and demographic data to predict MASLD diagnosis as defined by a set of ICD codes related to the disease. The model’s predicted probability of MASLD, here termed the in-silico MASLD (I-MASLD) score, provides a continuous risk estimate for each individual and serves as the phenotype in GWAS. This approach enables association testing in the absence of imaging-based liver phenotypes and facilitates the identification of genetic variants linked to subclinical or underdiagnosed disease. Using this framework, we recapitulate known MASLD-associated loci, including *PNPLA3*, *TM6SF2*, and the apolipoprotein A5 (*APOA5*) gene, and identify potentially novel associations, such as variants in the angiopoietin-like 4 (*ANGPTL4*) gene, that significantly increase MASLD risk. We also identify *APOA5* as a novel candidate against lean MASLD. These results highlight the utility of ML-derived phenotypes in enhancing genetic discovery in large-scale cohorts where conventional case/control labels are limited or imprecise.

## 2. Results

### 2.1. A Random Forest Model Accurately Predicts MASLD in Individuals

We trained a random forest classifier to predict MASLD status using 17 non-genomic clinical features, including laboratory measurements, vital signs, and demographic information (see Methods). MASLD cases were matched to controls based on age and body mass index (BMI) to minimize confounding prior to model training. The dataset was randomly divided into a training set (80%) and a test set (20%), with model development and evaluation performed separately to prevent overfitting. The classifier was trained to predict the presence of any of six ICD codes used as proxies for MASLD (see Methods). To reduce potential confounding from other liver diseases, individuals with corresponding ICD codes or with abnormal serum aspartate aminotransferase (AST) or alanine aminotransferase (ALT) levels were excluded from the control cohort, as elevated AST/ALT is indicative of hepatocellular injury likely due to underlying liver conditions (14).

For each individual, the model generates a probability score representing the predicted likelihood of MASLD. We term this predicted probability the in-silico MASLD (I-MASLD) score, a continuous, ML-derived estimate of disease risk based exclusively on clinical and demographic features.

The model demonstrated strong discriminative performance on the held-out test set. As shown in Figure 2a, the ROC curve indicates clear separation between MASLD cases and controls. The I-MASLD scores achieved comparable accuracy in both overweight (held out test set AUROC = 0.887) and lean (held out test set AUROC = 0.885) individuals. We further evaluated the predictive performance of the full model relative to each input feature individually. In Figure 2b, the AUROC of the I-MASLD score is plotted alongside the AUROCs of each clinical feature used as a univariate predictor, with each feature assessed independently against the binary MASLD label. Although certain liver enzymes, such as AST and ALT, as well as glucose levels, exhibited moderate predictive ability alone, the I-MASLD score consistently outperformed any single feature. These results indicate that the model effectively integrates partially informative signals across multiple features to generate a more robust overall risk estimate.

**Figure 1:**
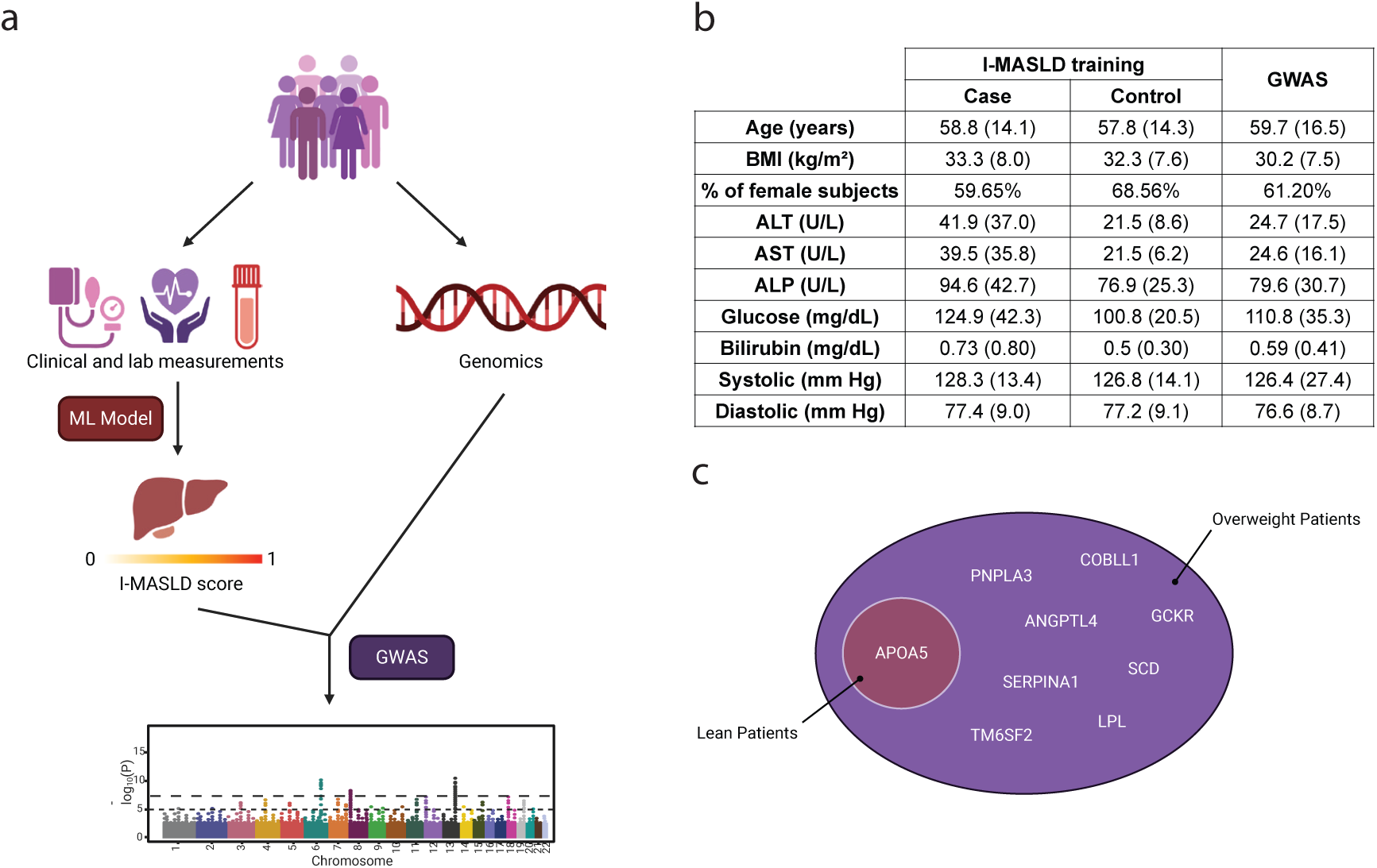
Overview of the ML-guided GWAS framework for MASLD. **(a)** Schematic of the machine-learning–guided genome-wide association framework. A random-forest model trained on clinical laboratory measurements, vital signs, and demographic variables predicts an in silico MASLD (I-MASLD) score representing each participant’s probability of MASLD. The continuous I-MASLD score is then used as a phenotype in GWAS of whole-exome sequencing variants from the All of Us cohort. **(b)** Summary of demographic and clinical characteristics of participants included in the I-MASLD model training and GWAS. Variables shown describe the cohort, not all model inputs. **(c)** Genome-wide association results using the I-MASLD score as a quantitative phenotype. Most loci were associated with MASLD in the context of elevated BMI, whereas variants in APOA5 showed strong association even among lean participants, suggesting a genetic risk component inde-pendent of overweight status.

**Figure 2:**
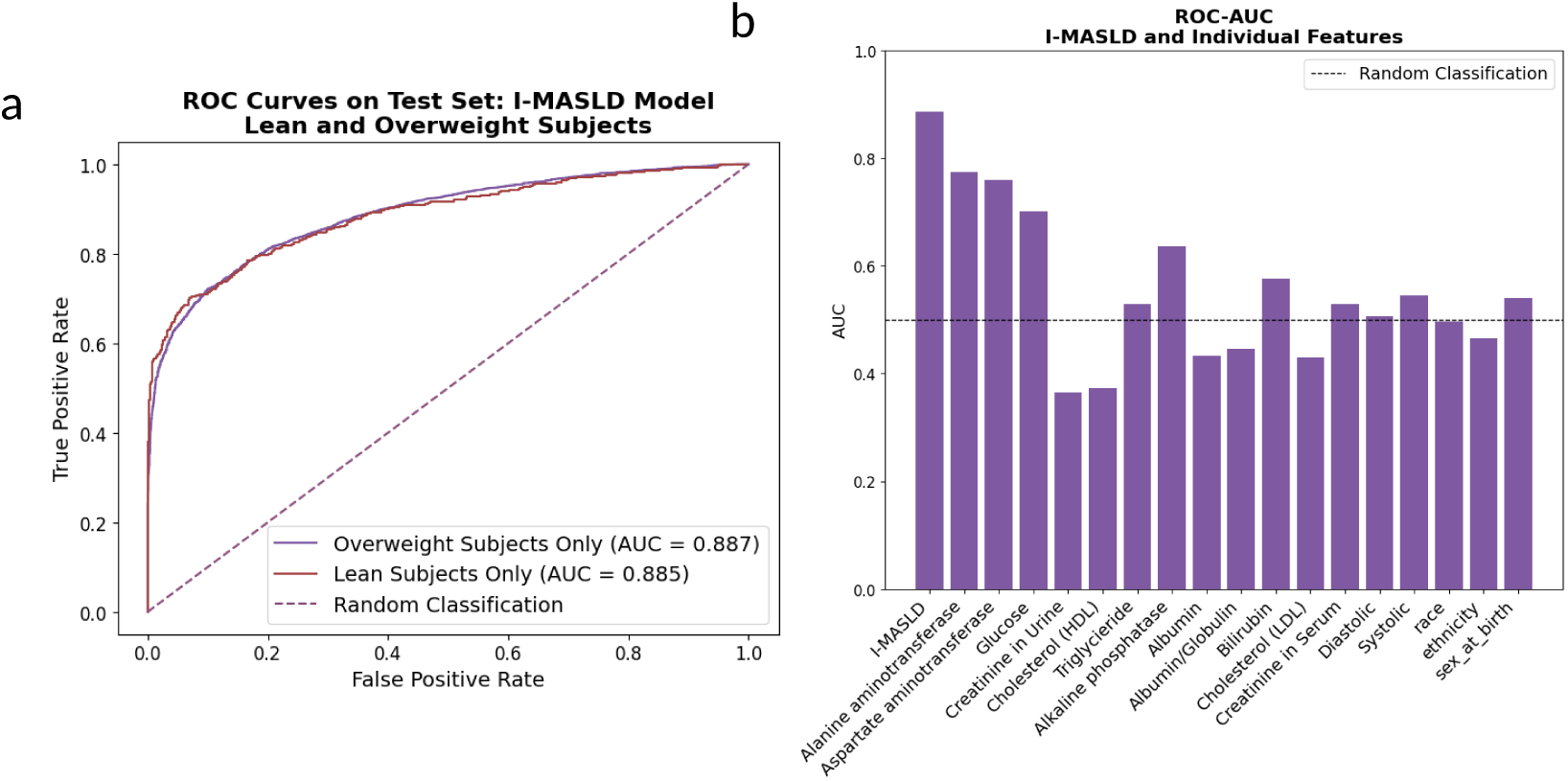
Performance of the I-MASLD model. **(a)** ROC curves for the set of overweight and lean subjects indicate clear seperation of MASLD cases and controls. **(b)** AUROC values for the full I-MASLD model and each input feature used individually as a univariate predictor. The I-MASLD score outperforms all single features, highlighting the benefit of integrating multiple clinical variables.

To assess which features contributed most to model predictions, we examined feature importances calculated using the Gini importance metric. In a random forest, Gini importance reflects the total reduction in impurity (measured by the Gini index) attributable to each feature across all decision trees in the ensemble. Features that frequently appear near the top of the trees and produce substantial improvements in classification accuracy are assigned higher importance scores, providing a heuristic measure of each feature’s contribution to the model’s decision-making process (15). As shown in Figure 3a, the highest-ranked features were clinically relevant laboratory measurements. The strongest contributors included AST, ALT and glucose, whereas other laboratory values, such as bilirubin and albumin/globulin ratio, had minimal influence on the predicted I-MASLD scores. Demographic variables, including race, ethnicity and sex assigned at birth also exhibited relatively low importance scores.

**Figure 3:**
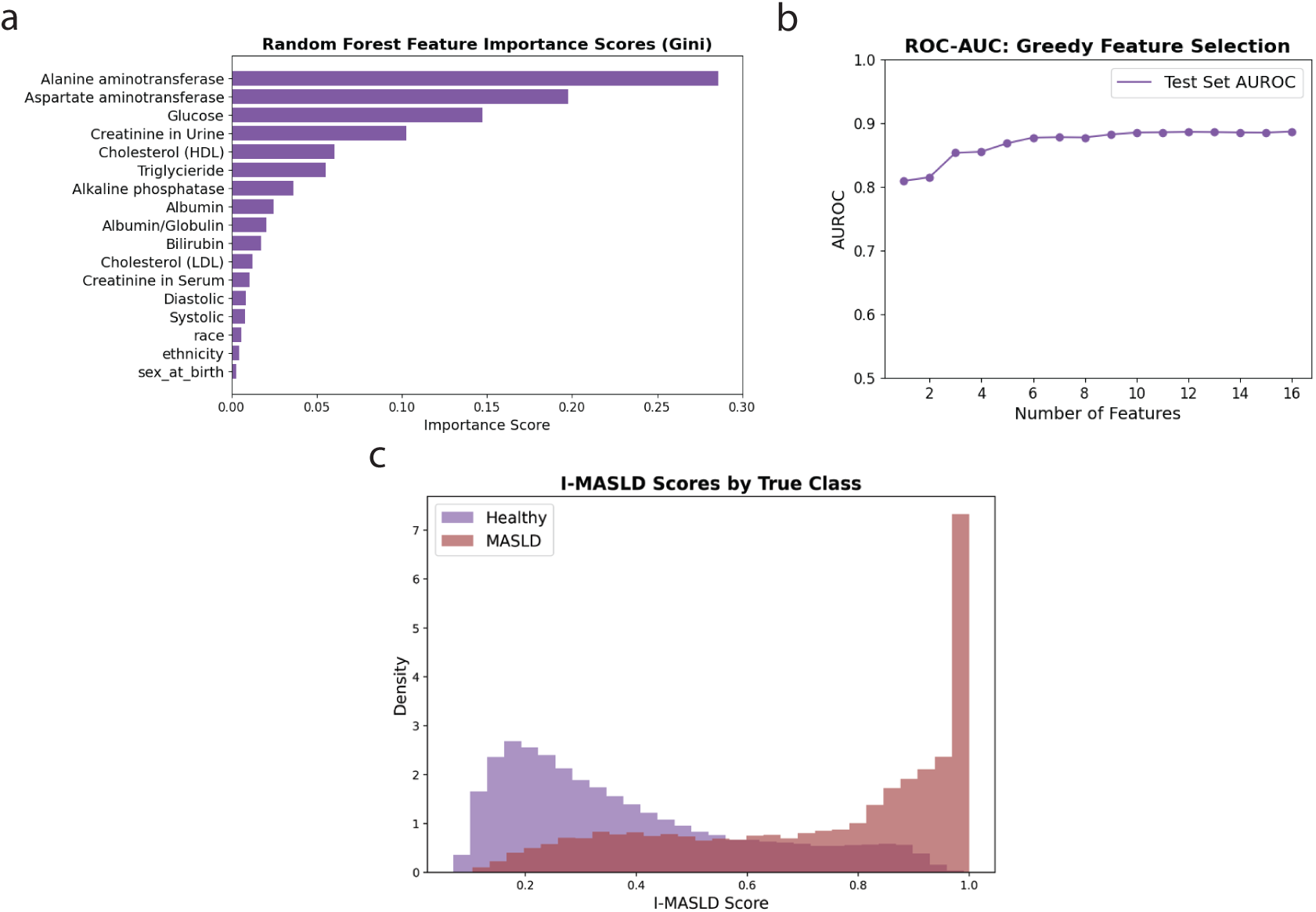
Interpretability and distribution of I-MASLD model predictions. **(a)** Gini feature importance scores from the trained random forest model. **(b)** AUROC curve for greedy feature selection, where features are added sequentially in order of importance. Performance increases rapidly with the first few features and plateaus after approximately 5 features, suggesting that most predictive value is captured by a core clinical subset. **(b)** Histogram of predicted I-MASLD scores for all individuals in the dataset. Individuals with a documented MASLD diagnosis show a right-skewed distribution while a broader distribution is observed among undiagnosed individuals.

To further quantify the cumulative effect of these features, we performed greedy feature selection by sequentially training models that added the next most important feature, based on Gini importance, to the existing feature set. As shown in Figure 3b, AUROC increased rapidly with the inclusion of the first few features and began to plateau after the top six variables, indicating that the I-MASLD score could be predicted with nearly equivalent accuracy using only AST, ALT, glucose, creatinine, triglycerides, and HDL cholesterol. The prominent importance of AST and ALT is consistent with their established role as liver enzymes commonly employed in initial MASLD screening (16; 17; 18; 19; 20). Notably, model performance did not decline substantially with the addition of further features, suggesting that lower-ranked variables were not detrimental. These results indicate that a small subset of clinical biomarkers captures the majority of the model’s discriminative power, while the remaining features contribute marginally but consistently.

To assess how the model stratifies risk across the full population, we plotted the distribution of I-MASLD scores for all individuals, including the training and test sets, those excluded from either set to balance the dataset, and subjects diagnosed with other liver-related diseases that were excluded during model training (figure 3c). Among individuals with a documented MASLD diagnosis, predicted scores were strongly right-skewed, with most individuals assigned high I-MASLD values. This pronounced skew indicates that the model is highly sensitive, correctly assigning elevated risk to nearly all known cases and thereby minimizing false negatives. A substantial proportion of apparently healthy subjects also exhibited I-MASLD scores above 0.5, suggesting elevated MASLD susceptibility and the potential presence of undiagnosed disease within this cohort. We further examined I-MASLD scores for control subjects who met the ICD exclusion criteria but had AST or ALT levels outside the normal range (Supplementary Figure 1). The predicted scores for these individuals were sharply right-skewed, indicating that most are likely to have MASLD. Notably, the small left-hand peak corresponded to subjects with AST and ALT below the normal range, whereas the larger right-hand peak comprised individuals with elevated AST and ALT, consistent with hepatocellular injury and potential undiagnosed MASLD cases.

### 2.2. The I-MASLD Score Generated by the Random Forest Allows for the Identification of Genetic Variants Associated with MASLD

Using whole-genome sequencing data, we performed GWAS with the Regenie (v4.1) (21) pipeline on exonic variants, analyzing overweight (*BMI >* 25 *kg/m*^2^) and lean (*BMI ≤* 25 *kg/m*^2^) individuals separately. Covariates included age, BMI, sex assigned at birth, presence of diabetes, dyslipidemia, hypertension (22), and genomic principal components to minimize potential confounding. Manhattan plots depicting association results for overweight and lean MASLD are shown in Figures 4a and c, respectively, while the corresponding quantile-quantile (QQ) plots (Figure 4b and c) indicated no evidence of systematic inflation in p-values. The genes corresponding to the identified loci are listed in Tables 1a and 1b for overweight and lean individuals respectively. Locus zoom plots visualize manhattan plots in the vicinity of each identified locus, and are also included in the Supplementary Materials. Sex chromosomes were excluded from the GWAS due to their distinct inheritance patterns, differences in ploidy between sexes, and complex dosage compensation mechanisms, which complicate association testing and can bias results if analyzed alongside autosomes.

**Figure 4:**
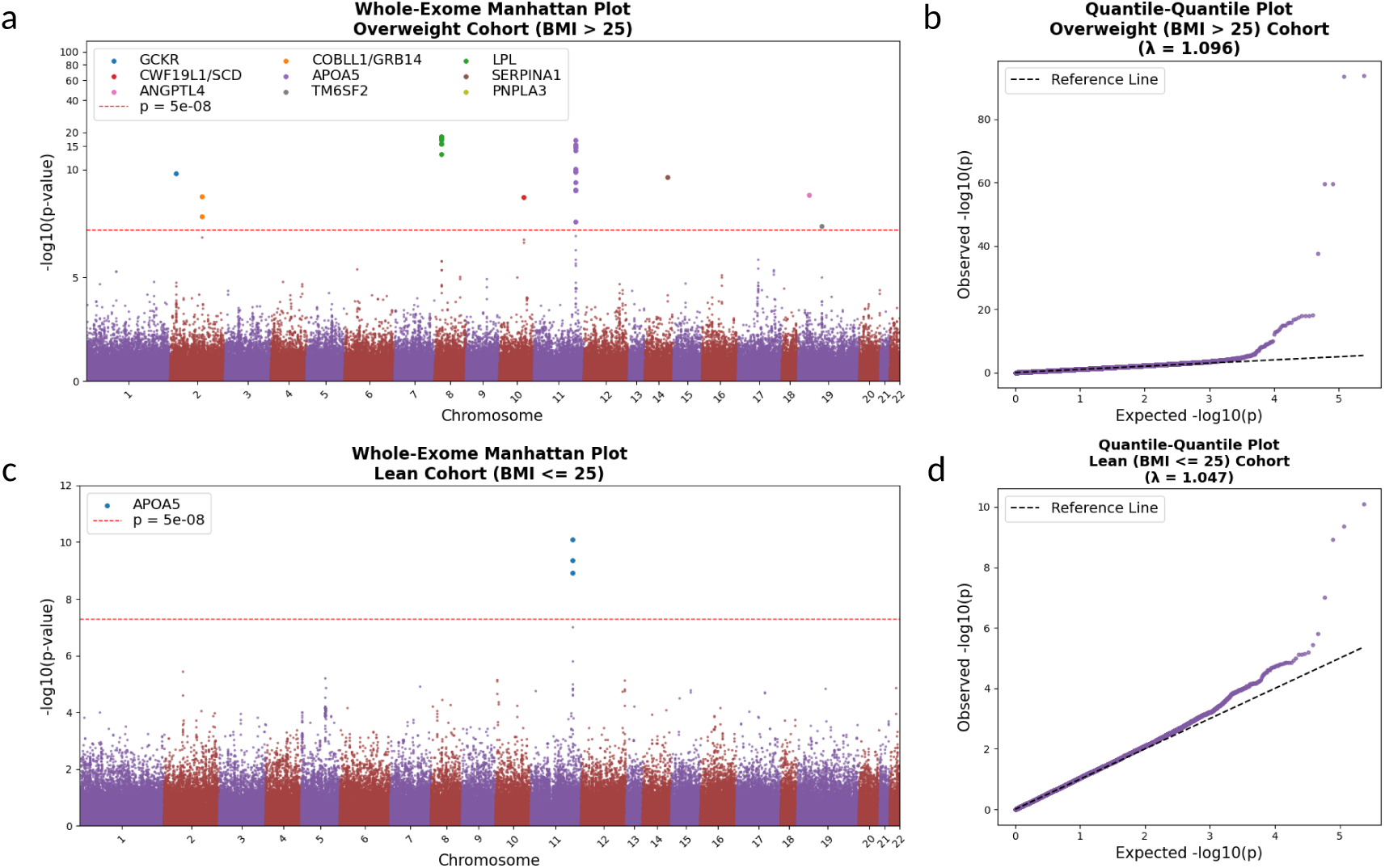
**(a), (b)**: Manhattan plot showing genome-wide association results using I-MASLD scores as a continuous phenotype for **(a)** overweight and **(b)** lean individuals. The genes identified as causal by the automated literature search as well as the genes corresponding to the SNPs that crossed the significance threshold are indicated in the plot legend.

**Table 1:**
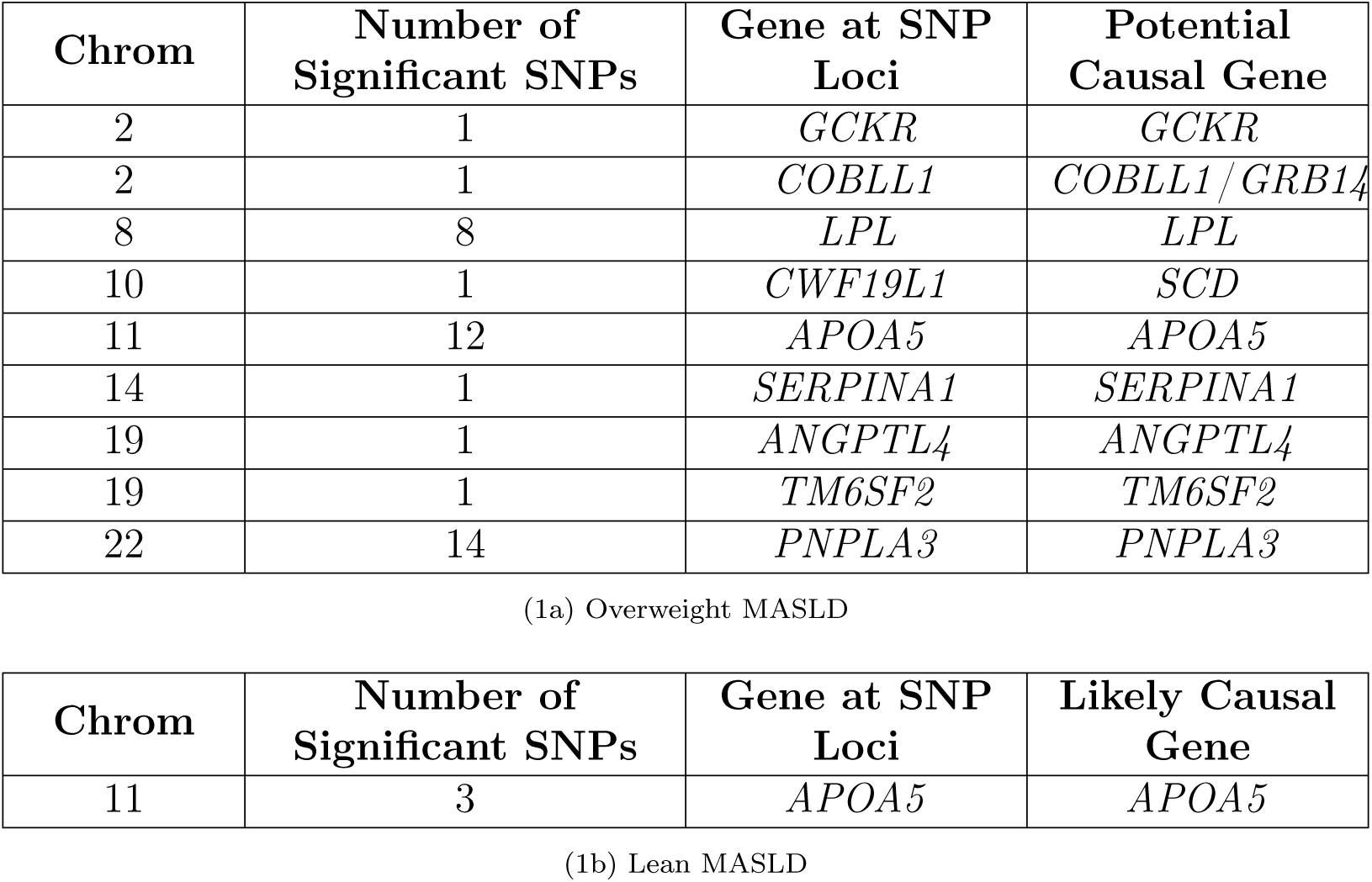
Genes which the significant SNPs for overweight and lean MASLD (Manhattan plots in figures 4a and b map to. Both, the gene containing the SNP locus and the LLM-suggested causal gene are listed.

Following the approach of Shringarpure et al. (23), we identified genes within 500 kb of loci surpassing the genome-wide significance threshold (5 *×* 10^−8^) and used an automated literature-synthesis process to aid causal-gene prioritization. This generated ranked candidate genes with brief justifications, and the results were reviewed for biological plausibility. In nearly all cases, the top candidate corresponded to the gene containing the lead SNP; the only ambiguous locus was on chromosome 2, where both *COBLL1* and *GRB14*, adjacent genes previously linked to MASLD, were supported (Supplementary Figure 2).

For MASLD in overweight individuals, several peaks were observed in the Manhattan plot, some of which contained SNPs exceeding the genome-wide significance threshold. The first significant locus on chromosome 2 maps to the glucokinase regulator (*GCKR*) gene. Variants in *GCKR* have been associated with elevated triglycerides, increased hepatic fat content, and MASLD susceptibility in multiple population-based studies (24). Functional studies in humans and mouse models indicate that *GCKR* modulates hepatic glucose and lipid metabolism, promoting de novo lipogenesis and contributing to steatosis when dysregulated (25).

The second significant locus on chromosome 2 contains variants within the cordonbleu WH2 repeat protein like 1 (*COBLL1*) gene. *COBLL1* influences lipid metabolism and is a key regulator of lipid homeostasis, in addition to modulating cellular growth and apoptosis (26; 27). SNPs in *COBLL1* have been previously associated with MASLD and related metabolic traits in multiple GWAS (28; 29; 30). A nearby gene, growth factor receptor-bound protein 14 (*GRB14*), located within the same locus, also represents a plausible causal candidate. *GRB14* encodes an adaptor protein that negatively regulates insulin signaling in hepatocytes, and its dysregulation has been linked to increased hepatic lipid accumulation and steatosis in both human cohorts and animal models (29). Given their proximity and shared functional relevance to hepatic lipid metabolism, either *COBLL1* or *GRB14* may underlie the observed association.

Several loci on chromosome 8, mapping to the lipoprotein lipase (*LPL*) gene, were also highly significant. *LPL* is crucial for hydrolyzing triglycerides in the blood, playing a central role in lipid metabolism and VLDL synthesis and secretion. Specific SNPs within *LPL* have been associated with variations in triglycerides, total cholesterol, and LDL levels (31), all of which are implicated in MASLD (32). Although studies directly linking *LPL* variants to MASLD progression are still emerging, its central role in lipid metabolism makes it a key gene for understanding MASLD, particularly regarding its impact on lipid profiles.

The significant locus on chromosome 10 contains a variant within the CWF19-like cell cycle control factor 1 (*CWF19L1*) gene, part of the *ERLIN1* –*CHUK* –*CWF19L1* cluster previously associated with hepatic fat accumulation and inflammation in a genome-wide association study (33). The recurrence of this signal in our analysis, despite its limited recognition in more recent MASLD studies, suggests that this region may warrant renewed investigation to clarify its role in hepatic lipid metabolism and disease susceptibility. A nearby gene, stearoyl-CoA desaturase (*SCD*), located approximately 100 kb from the lead SNP, represents another biologically plausible causal candidate. *SCD* encodes an enzyme that catalyzes the synthesis of monounsaturated fatty acids (MUFAs) from saturated fatty acids, a key step in lipid biosynthesis. Overproduction of MUFAs can promote hepatic fat accumulation and steatosis (34), and elevated *SCD* expression has been reported in MASLD patients (35), suggesting that gain-of-function variants may contribute to disease onset through enhanced de novo lipogenesis.

On chromosome 11, several significant variants are found in the *BUD13* -*ZPR1* - *APOA5* region. Among these, *APOA5*, which regulates triglyceride metabolism by modulating enzymes involved in the degradation of circulating triglycerides (36), represents the most biologically plausible causal gene. *APOA5* contributes to the formation and stability of chylomicrons and VLDLs, which transport triglycerides from the liver and intestines to peripheral tissues (37). Prior studies indicate that *APOA5* overexpression promotes MASLD by impairing triglyceride clearance, resulting in hepatocellular lipid accumulation (38), and it has been proposed as a target for interventions such as olazipine (39).

The SNP surpassing the significance threshold on chromosome 14 corresponds to the Z-allele variant of the SERPIN-family A member 1 (*SERPINA1*) gene, which has been strongly associated with MASLD (40). *SERPINA1*, also known as alpha-1 antitrypsin (*A1AT*), is a serine protease inhibitor primarily produced by hepatocytes. Misfolded variants of *A1AT* accumulate in the hepatocyte endoplasmic reticulum, leading to liver injury, inflammation, and fibrosis (41).

Variants mapping to two distinct genes on chromosome 19, *TM6SF2* and *ANGPTL4*, were identified. *TM6SF2* regulates hepatic lipid accumulation by modulating triglyceride secretion and plays a key role in maintaining cholesterol homeostasis (42; 43). The role of *TM6SF2* in MASLD and related cardiometabolic disorders is well established, with several of its variants previously associated with increased disease risk (44; 45; 46). *ANGPTL4*, an inhibitor of *LPL*, regulates triglyceride hydrolysis and the delivery of fatty acids to adipose tissue. Altered *ANGPTL4* function disrupts *LPL*-mediated triglyceride metabolism and hepatic lipid uptake, potentially contributing to MASLD pathogenesis (47). Furthermore, depletion of *ANGPTL4* in mouse models has been shown to exert a protective effect against hepatic steatosis (48). Notably, this variant was not present in any existing SNP databases, indicating the discovery of a potentially novel and clinically relevant variant.

The significant loci on chromosome 22 represent the replication of several previously reported variants within the *PNPLA3* and sorting and assembly machinery component 50 homolog (*SAMM50*) gene cluster. *PNPLA3* encodes a triglyceride lipase that mediates triacylglycerol hydrolysis in adipocytes (49) and is identified as the causal gene. Furthermore, mutations in *PNPLA3* have been shown to be associated with MASLD in several studies, and it is the most replicated genetic risk factor for MASLD across multiple GWAS studies (6; 10; 50).

For MASLD in lean individuals, we identified three variants within the *APOA5* gene that were significantly associated with disease risk. Notably, these variants were also significant in the overweight MASLD cohort, indicating shared genetic determinants across BMI strata. However, their association with MASLD among lean individuals suggests that APOA5 variants exert sufficiently strong effects on hepatic lipid metabolism to confer disease risk even in the absence of obesity. Previous studies investigating the genetic architecture of lean MASLD have primarily implicated variants in *PNPLA3*, *GCKR*, *SAMM50*, and the SURP and G-patch domain containing (*SUGP1*) gene (22; 51; 52). The identification of *APOA5* variants in this study, to the best of our knowledge, represents a novel genetic locus associated with lean MASLD. Recent reviews of lean MASLD report that, although triglyceride levels in lean MASLD patients are lower than those in overweight MASLD patients, they remain elevated compared to lean or overweight individuals with healthy liver function (53). This observation, together with our discovery of *APOA5* variants, may suggest that MASLD onset in lean individuals is driven by increased de novo lipogenesis resulting from elevated *APOA5* activity, leading to hepatocellular damage and inflammation. Other members of the apolipoprotein family, such as the apolipoprotein A1 (*APOA1*) gene, have also been implicated in lean MASLD (54), further underscoring the potential role of lipid transport and metabolism pathways in the disease’s pathogenesis.

## 3. Discussion

In this study, we implemented a machine learning–guided GWAS framework to identify genetic risk factors for MASLD in individuals with obesity. Using clinical and laboratory features from the *All of Us* Research Program, we trained a random forest classifier to derive a continuous in silico MASLD (I-MASLD) score that reflects an individual’s likelihood of disease. This ML-derived quantitative phenotype was then used as a trait in GWAS, serving as a proxy for MASLD and enabling the discovery of genetic associations beyond those detectable through conventional case–control definitions.

In the cohort of overweight individuals, this approach successfully identified new risk alleles in *LPL* and *ANGPTL4*, genes with well-characterized roles in lipid metabolism but limited prior evidence linking them directly to MASLD pathogenesis. The appearance of both LPL and ANGPTL4, two key regulators of triglyceride clearance and lipoprotein lipase activity, reinforces the involvement of lipid metabolism in MASLD. Although LPL has been extensively studied in the context of plasma lipid levels and cardiometabolic traits, direct associations with MASLD are only beginning to emerge, making its recurrence in our analysis notable (31). ANGPTL4, an inhibitor of LPL that regulates plasma triglyceride levels during fasting and metabolic stress, has an even more limited publication record in MASLD genetics (55). Despite clear mechanistic links to lipid handling, a recent clinical review emphasizes that the impact of ANGPTL4 on MASLD remains unresolved and has not been demonstrated in human genetic studies (55). Our finding therefore provides new empirical support for a role of ANGPTL4 in MASLD susceptibility. Notably, the lead variant we observe appears to be absent from dbSNP, suggesting that it may represent a previously unreported allele with potential clinical relevance. Together, these observations highlight ANGPTL4 as a promising candidate for follow-up studies aimed at understanding how disruptions in triglyceride regulation contribute to hepatic fat accumulation. Additionally, we recovered established MASLD risk loci, including *PNPLA3*, *GCKR*, and *TM6SF2*.

Recent studies on MASLD have looked beyond fat accumulation alone, and have emphasized the role of inflammation in the onset and progression of MASLD (56; 57; 58). In line with this, our GWAS identified associations in *SERPINA1* and *CWF19L1* which have roles in driving inflammation. While the mechanism by which *SERPINA1* contributes to MASLD is well-characterized, *CWF19L1* remains poorly understood at a functional level. However, *CWF19L1* lies within the *ERLIN1–CHUK–CWF19L1* gene cluster, gene cluster, in which common variants have previously been shown to influence both CT-quantified liver fat and ALT levels (33). The recurrence of this locus in our MASLD GWAS, using an independent cohort and a distinct, ML-derived phenotype, strengthens the evidence that *ERLIN1–CHUK–CWF19L1* variation modulates susceptibility to steatotic liver disease and suggests that *CWF19L1* may contribute to hepatic and inflammatory responses through mechanisms that remain to be elucidated.

Finally, we propose *APOA5* as a novel candidate gene contributing to MASLD onset in lean individuals, potentially through enhanced de novo lipogenesis. The discovery of *APOA5* variants specifically associated with lean MASLD highlights a specific disease mechanism and warrants validation in targeted animal model studies to elucidate the underlying biological pathways. These findings underscore the potential of ML-guided phenotyping to uncover both known and previously unrecognized genetic mechanisms underlying MASLD.

The ML-guided GWAS framework addresses several challenges in performing GWAS studies on MASLD. First, the reliance on diagnostic codes such as ICD-10 K76.0 can be limiting in the context of MASLD, a condition known to be under-diagnosed and highly variable in clinical presentation. Second, the *All of Us* cohort, while uniquely diverse, does not currently contain imaging- or biopsy-based liver phenotypes for most participants. The I-MASLD score provides a scalable, data-driven alternative that leverages more readily available clinical and lab values to estimate disease risk across a broad population. Additionally, recent reports on GWAS of biobank-scale data support the use of engineered phenotypes rather than simple case-control labels (59). The observed alignment of GWAS hits with known biology further reinforces the clinical relevance of this derived phenotype.

The present findings open several avenues for further investigation. Although the I-MASLD score is designed to interpolate between case and control labels and can fill the gaps where individuals with severe MASLD are misdiagnosed, it remains trained on ICD-based labels, which are an imperfect proxy for true disease status. This reliance on structured diagnosis codes is not the optimal approach for defining ground truth and inevitably introduces noise into the training labels, even if the model can compensate for some of it. Ideally, the I-MASLD scores would be validated against imaging-based phenotypes such as MRI-PDFF. However, such data are not available in *All of Us*. Future studies on data from the UK Biobank may be able to fill this gap. Second, given the demographic diversity of the *All of Us* cohort, ancestry-stratified analyses could uncover population-specific genetic associations that may be missed in aggregate models and improve understanding of MASLD risk in historically understudied groups. Third, as lifestyle and dietary modifications remain the primary clinical interventions for MASLD, future studies should examine gene–environment interactions by integrating the SNPs identified through the I-MASLD–guided GWAS with data on dietary intake, physical activity, and other environmental exposures. Such analyses could clarify how genetic susceptibility interacts with modifiable behavioral factors to shape disease risk. Finally, developing interpretable ML frameworks to model these nonlinear interactions represents a promising direction for gaining a more holistic view of MASLD etiology and informing personalized prevention strategies.

## 4. Methods

### 4.1. Data

We used data from the *All of Us* Research Program Registered Tier v8, which includes electronic health records (EHR), laboratory test results, physical measurements, and self-reported demographic information (8). Analyses were restricted to 414,830 participants with available whole genome sequencing (WGS) data. MASLD cases were defined as individuals diagnosed with at least one of the six ICD codes listed in table 2. To avoid including individuals with possible undiagnosed liver disease, we excluded from the control group all subjects with abnormal serum AST or ALT levels, as these are indicative of liver injury. The reference ranges used for AST were 8–42 U/L for females and 8–48 U/L for males, and for ALT were 7–45 U/L for females and 7–55 U/L for males.

**Table 2:**
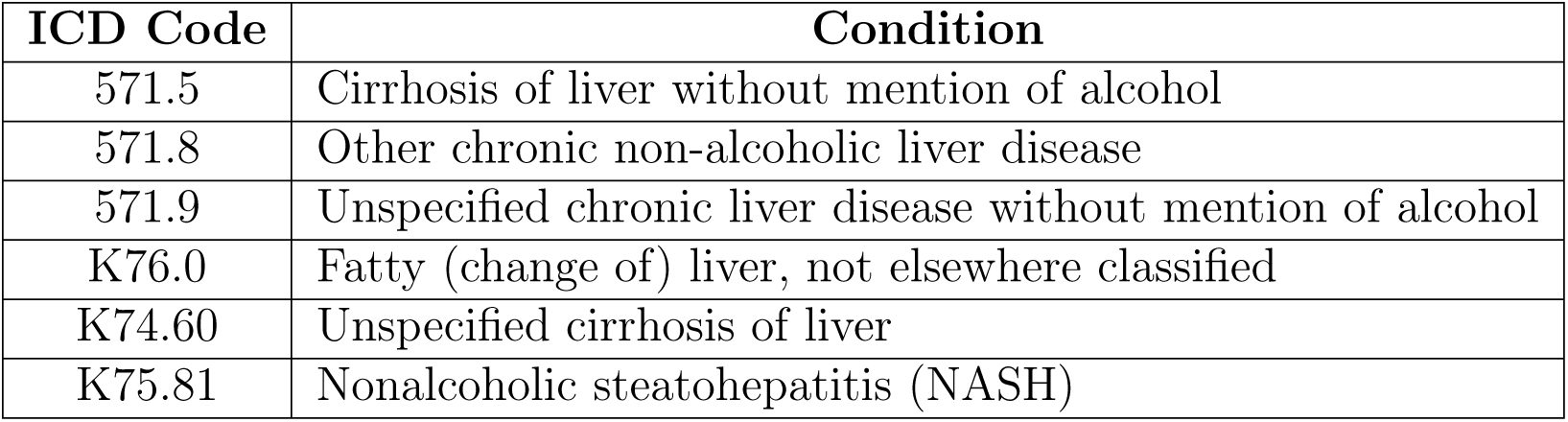
ICD codes used to identify subjects with MASLD in the *All of Us* dataset.

| ICD Code | Condition |
| --- | --- |
| 571.5 | Cirrhosis of liver without mention of alcohol |
| 571.8 | Other chronic non-alcoholic liver disease |
| 571.9 | Unspecified chronic liver disease without mention of alcohol |
| K76.0 | Fatty (change of) liver, not elsewhere classified |
| K74.60 | Unspecified cirrhosis of liver |
| K75.81 | Nonalcoholic steatohepatitis (NASH) |

We observed that the BMI values reported within the *All of Us* dataset occasionally deviated from those calculated directly from height and weight measurements (see supplementary figure 2). Therefore, we used the calculated BMI values for all downstream analyses after averaging all available height and weight readings. Individuals with implausible height (below 1.0 m or above 2.4 m) or weight (below 10 kg or above 800 kg) were removed from both case and control groups.

To train the MASLD prediction model, we excluded from the control pool any subjects diagnosed with other liver-related conditions (listed in supplementary table 2) to minimize confounding. The resulting dataset used for model training comprised 26,062 MASLD cases and 62,786 controls. To construct a balanced training cohort, we matched MASLD cases to healthy controls by age (±1 year) and BMI (±2 *kg/m*^2^) using a greedy nearest-neighbor matching approach. Matching was applied exclusively for model training and not for downstream genome-wide association analyses.

The machine learning model was trained on 17 input features: 12 laboratory measurements, two vital signs and three demographic variables as listed in supplementary table 1. Individuals missing more than 11 of these features were excluded (supplementary figure 4), resulting in the removal of 748 case subjects and no controls. Remaining missing values were imputed using mean imputation for continuous variables and most-frequent-value imputation for categorical variables.

The dataset reflects the demographic diversity of the *All of Us* cohort. As shown in supplementary figure 3, approximately 53% of participants identified as White, 15% as Black or African American, 3% as Asian, 1.6% as American Indian or Alaska Native, and 0.7% as Asian, Middle Eastern, or Pacific Islander; 22% did not report race, and 4.5% reported more than one race. For ethnicity, 75% identified as not Hispanic or Latino, 23% as Hispanic or Latino, and 2% did not report ethnicity. This diversity enables the modeling of MASLD risk across a broad range of ancestries and demographic backgrounds.

### 4.2. I-MASLD Score Generation

To generate a continuous MASLD risk score for genome-wide association analysis, we trained a random forest classifier to predict MASLD diagnosis based on non-genomic clinical features. The training dataset consisted of an approximately 1:1 age- and BMI-matched case–control cohort, as described in the previous section.

The model incorporated 17 input features, including 12 laboratory measurements, 2 vital signs, and 4 demographic variables, as previously detailed. Prior to training, missing values in continuous variables were imputed using the column mean, while missing categorical values were imputed with the most frequent category. Categorical variables were then encoded as integers according to the number of unique categories present in the training set. All continuous features were standardized to have zero mean and unit variance.

The dataset was randomly split into training and testing subsets in an 80:20 ratio. A random forest classifier was trained using 100 decision trees, with bootstrap sampling enabled and feature subsampling at each split set to the square root of the total number of input features. Trees were allowed to grow to a maximum depth of 9, or until all terminal nodes were pure or contained fewer than two samples.

After training, the model was applied to the entire cohort of case and control individuals, including the control subjects excluded during training set balancing and those diagnosed with other MASLD-related liver conditions, irrespective of formal diagnosis status. For each individual, the model produced a predicted probability of MASLD, referred to as the *in-silico* MASLD (I-MASLD) score, a continuous, machine learning–derived estimate of disease risk used as the phenotype in subsequent genome-wide association analyses.

### 4.3. ML-Guided GWAS

We conducted a genome-wide association study (GWAS) to identify genetic variants associated with the I-MASLD score using whole genome sequencing (WGS) data from the *All of Us* Research Program (v8). The I-MASLD score, derived from non-genomic clinical features, was used as a continuous phenotype representing the predicted risk of MASLD. GWAS was performed separately for 128,717 overweight and 43,462 lean individuals, classified based on body mass index (BMI) calculated from height and weight measurements. Individuals with a BMI greater than 25*, kg/m*^2^ were categorized as overweight.

Variants across the whole exome were analyzed after filtering to retain only those with a minor allele frequency (MAF) *≥* 1. Association analysis was carried out using Regenie (v4.1) (21), which implements a two-step procedure for genome-wide association testing. In Step 1, a whole-genome regression model is fit using ridge regression within a cross-validation framework to capture polygenic effects and generate phenotype predictions while accounting for relatedness and population structure. In Step 2, single-variant association testing is performed using these predictions as covariates in a fast approximate mixed model, thereby controlling for confounding and enabling efficient large-scale scans.

The regression models included age, BMI, sex assigned at birth, and the first sixteen principal components (PCs) of genetic ancestry as covariates. These PCs, derived from common variants, were obtained from ancestry predictions provided by *All of Us* and were used to adjust for population stratification. Quantile–quantile (QQ) plots were generated to assess potential inflation or deflation of test statistics and to evaluate the overall calibration of association p-values.

## Data Availability

All data used in the study are available on the All of Us Workbench.

## 5. Data and Code Availability

The ML-driven GWAS pipeline for identifying genetic risk factors for MASLD is available within the All of Us Researcher Workbench (https://workbench.researchallofus.org/workspaces/aou-rw-919b8eca/mlguidedgwas/data).

## 6. Acknowledgements

We gratefully acknowledge *All of Us* participants for their contributions, without whom this research would not have been possible. We also thank the National Institutes of Health’s *All of Us* Research Program for making available the participant data examined in this study. AN and KK were supported by the Mayo Clinic and Illinois Alliance Fellowship for Technology-Based Healthcare. AN, SMD and SM received funding from the Personalized Nutrition Initiative (PNI) at the University of Illinois, and the PNI External Partners Directed Research Program for this work.

