## Supplemental Information for "ML-Guided GWAS Reveals Genetic Architectures for MASLD for Overweight and Lean Individuals in the *All of Us* Cohort"

---

### Supplementary Material

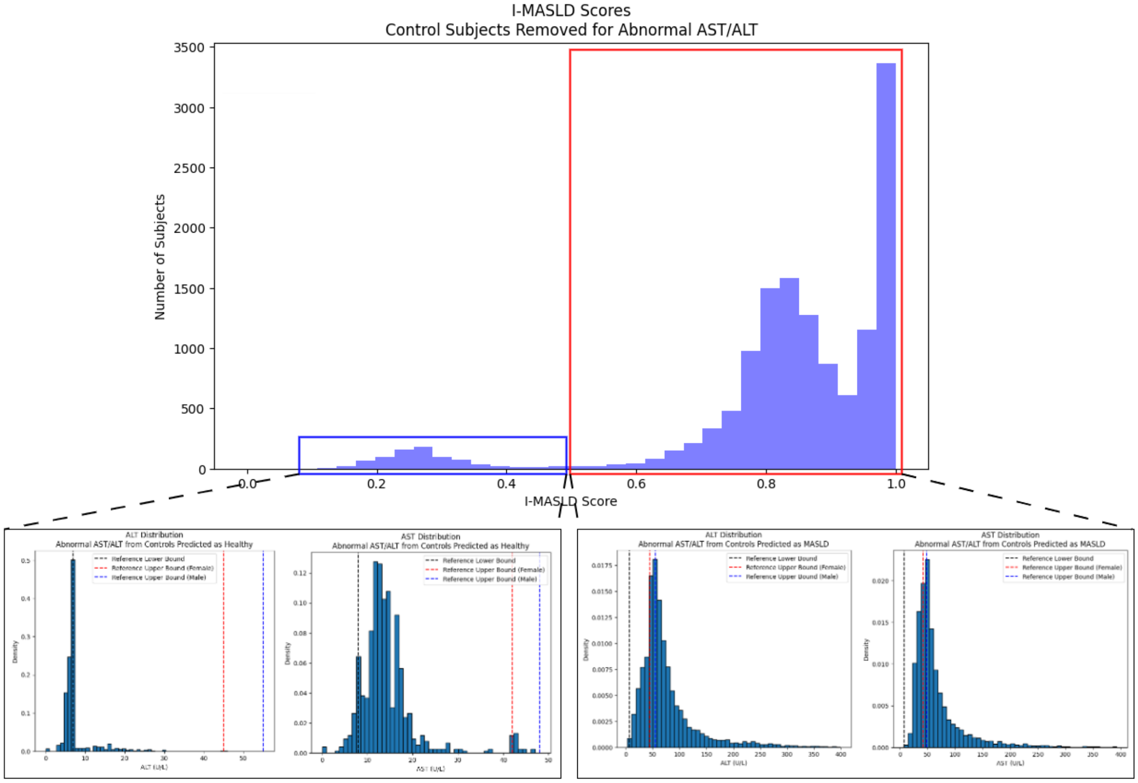


Figure 1: Distribution of I-MASLD scores for subjects with AST and ALT outside the reference range. The majority of these subjects have AST and ALT values above the reference range and are predicted as having MASLD. The small peak on the left representing healthy subjects contains subjects with AST and ALT below the reference range.

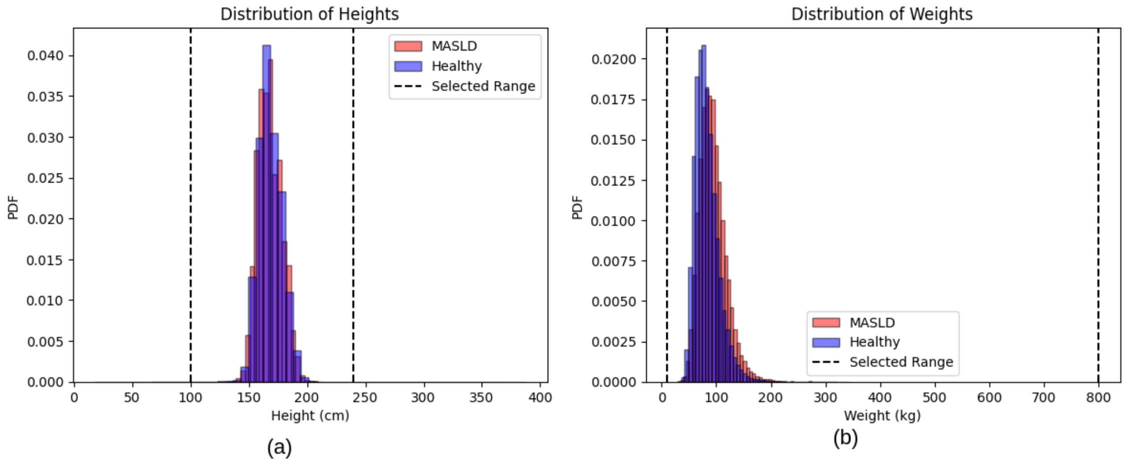


Figure 2: Height and weight distributions for subjects from the *All of Us* database under study along with the thresholds used for filtering.

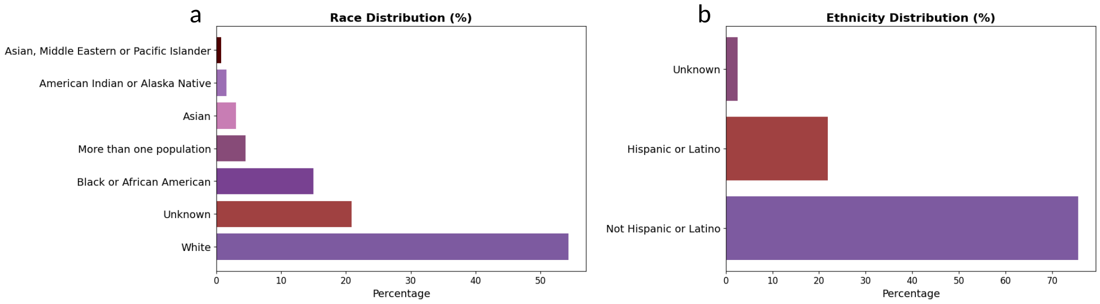


Figure 3: **Demographic diversity of the study population.** **(a)** Distribution of self-reported race and **(b)** ethnicity among individuals in this study from the *All of Us* cohort.

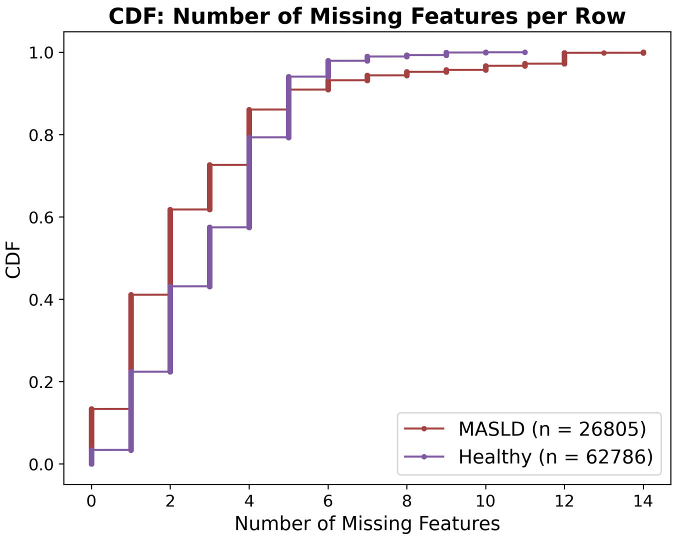


Figure 4: **Missing features in the dataset.** Cumulative distribution function (CDF) of the number of missing features per individual in the MASLD case (red) and control (blue) groups prior to imputation. Most individuals had few or no missing values across the 17 selected features, allowing for effective mean and mode imputation without the need for participant exclusion.

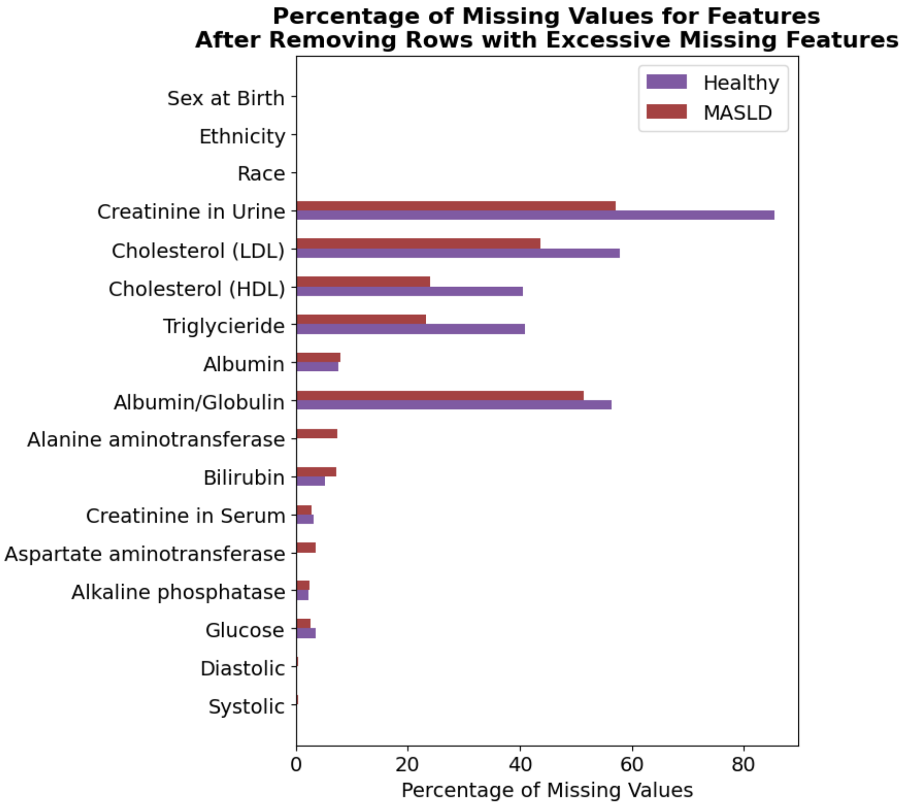


Figure 5: **Percentage of missing values per feature in the dataset after removing rows with excessive missing features.** Although some features had a high percentage of missing values, they did not contribute as significantly to the I-MASLD score predictions (see figures 3a and 3b in the main text).

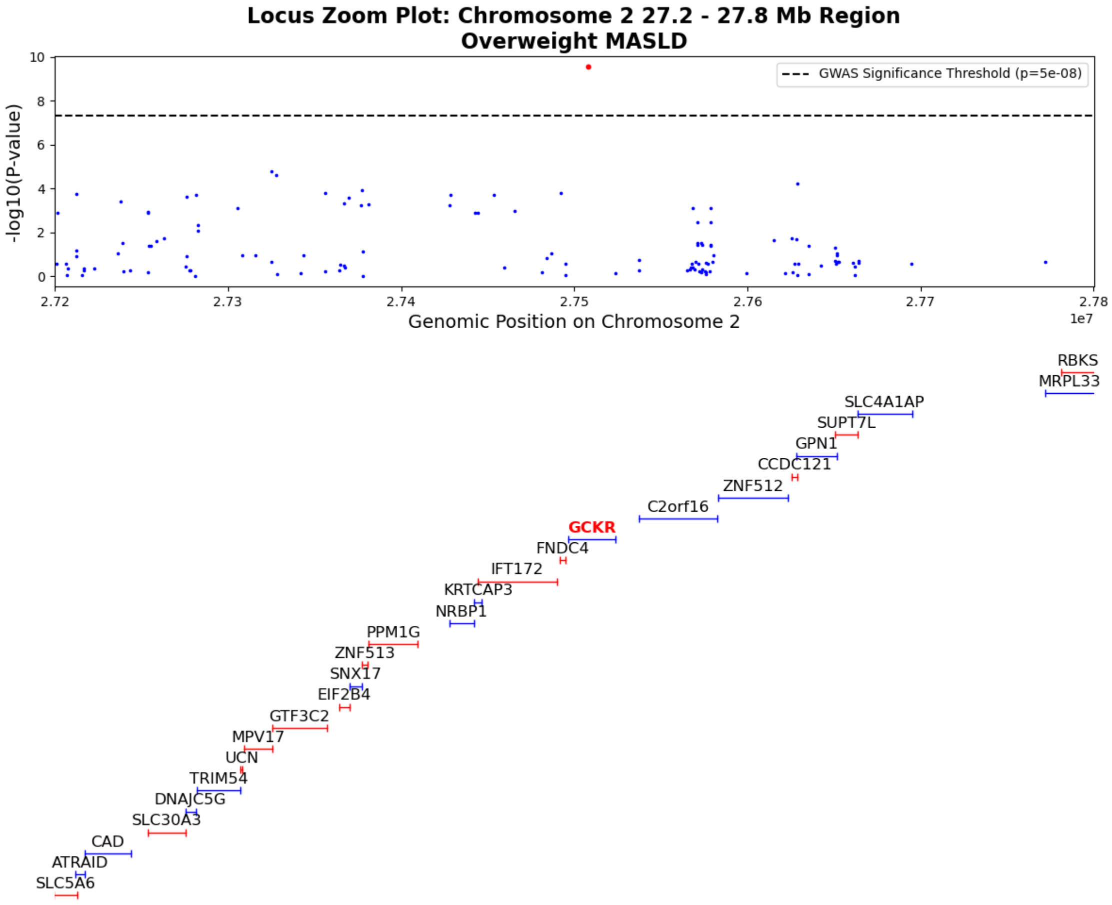


Figure 6: Locus zoom plot visualizing the p-values of significance for genetic variants for overweight MASLD along with genes near to the significant loci on chromosome 2 in the 27.2 to 27.8 Mb region. Genes containing significant loci are highlighted in bold and LLM-identified causal genes are colored red.

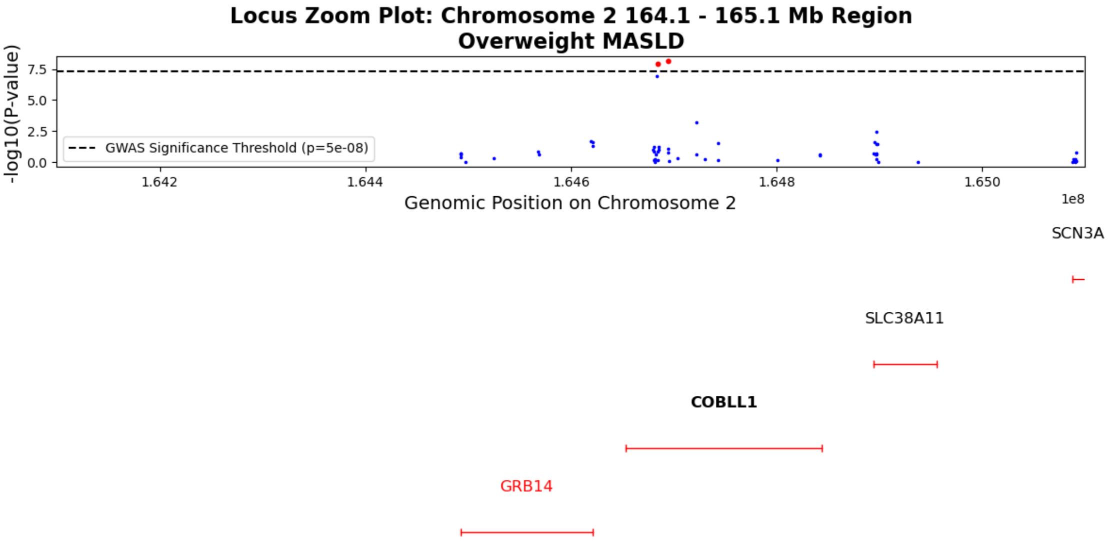


Figure 7: Locus zoom plot visualizing the p-values of significance for genetic variants for overweight MASLD along with genes near to the significant loci on chromosome 2 in the 164.1 to 165.1 Mb region. Genes containing significant loci are highlighted in bold and LLM-identified causal genes are colored red.

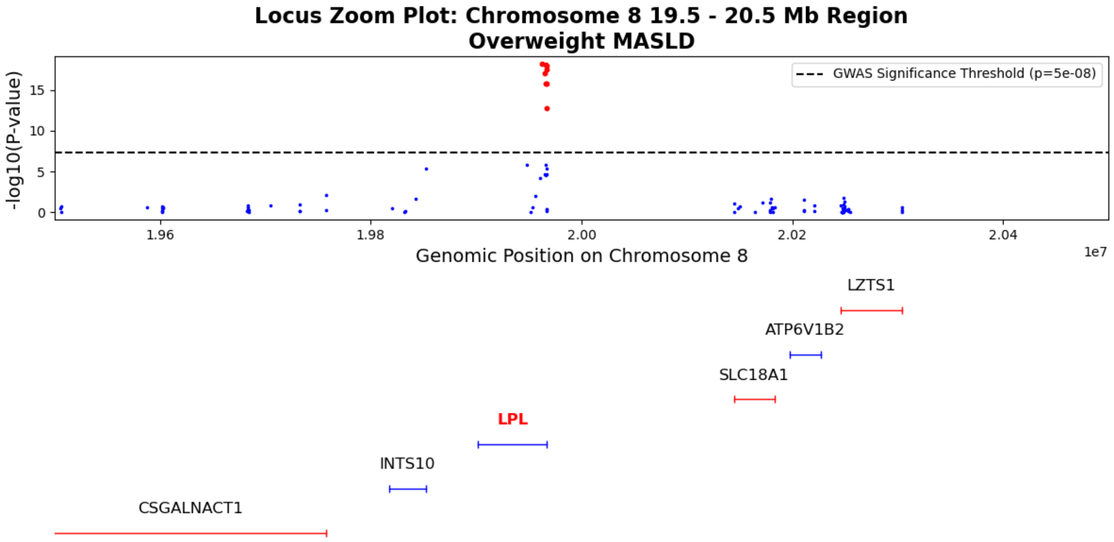


Figure 8: Locus zoom plot visualizing the p-values of significance for genetic variants for overweight MASLD along with genes near to the significant loci on chromosome 8 in the 19.5 to 20.5 Mb region. Genes containing significant loci are highlighted in bold and LLM-identified causal genes are colored red.

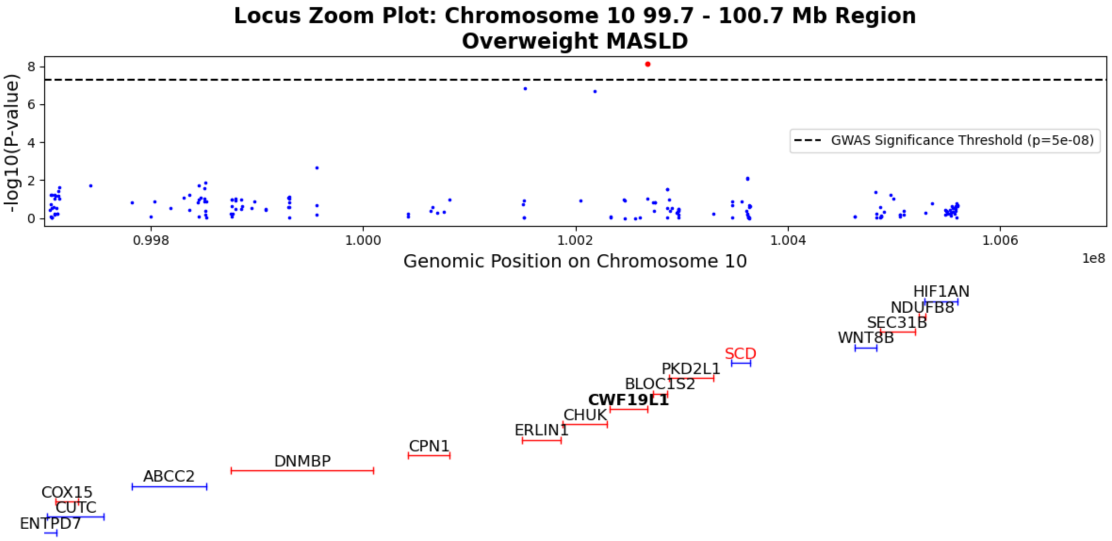


Figure 9: Locus zoom plot visualizing the p-values of significance for genetic variants for overweight MASLD along with genes near to the significant loci on chromosome 10 in the 99.7 to 100.7 Mb region. Genes containing significant loci are highlighted in bold and LLM-identified causal genes are colored red.

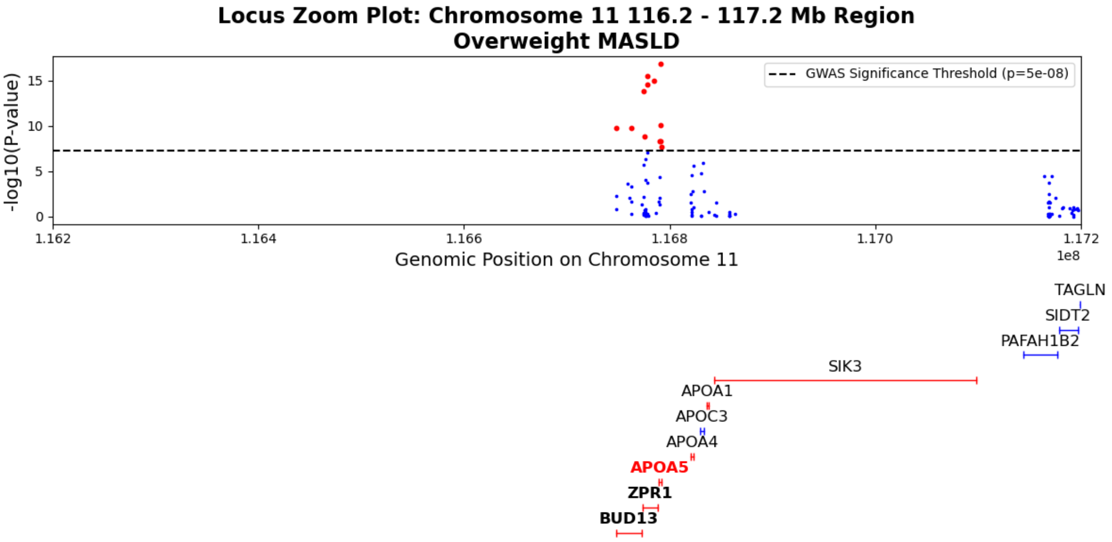


Figure 10: Locus zoom plot visualizing the p-values of significance for genetic variants for overweight MASLD along with genes near to the significant loci on chromosome 11 in the 116.2 to 117.2 Mb region. Genes containing significant loci are highlighted in bold and LLM-identified causal genes are colored red.

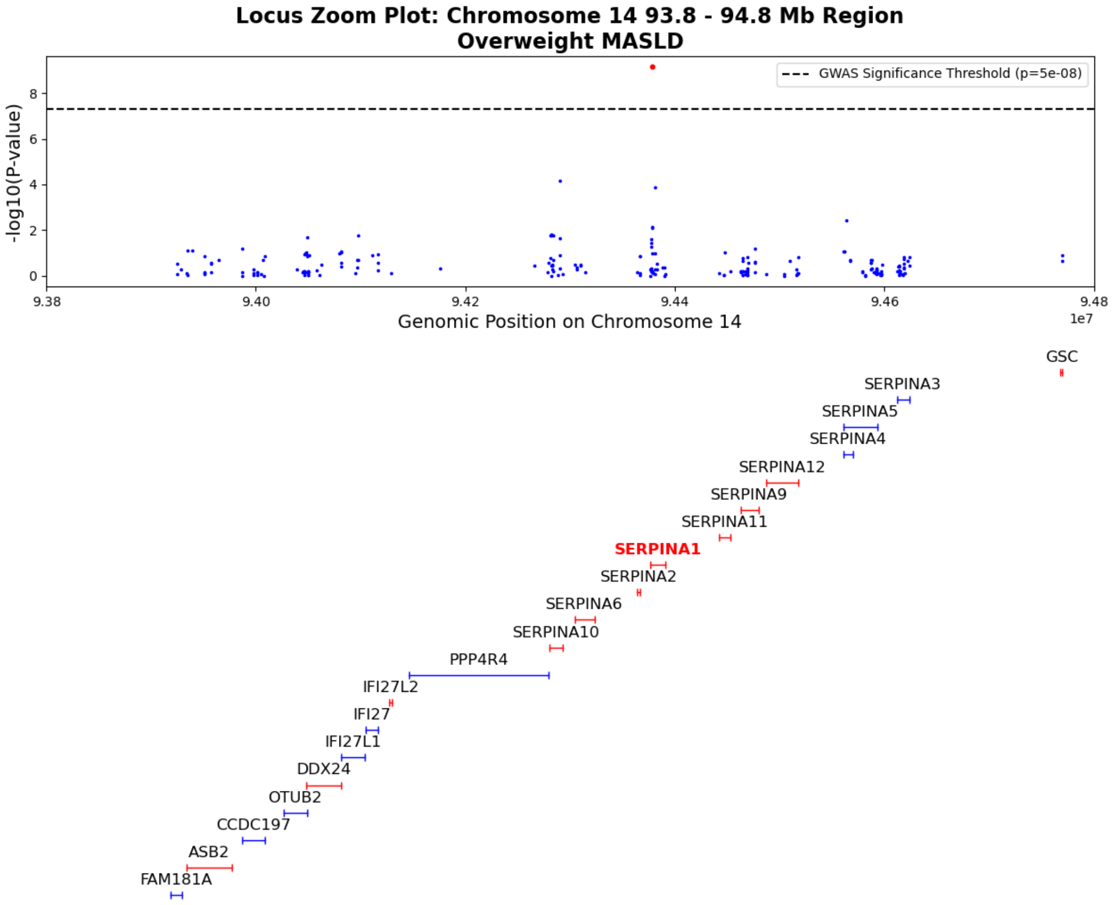


Figure 11: Locus zoom plot visualizing the p-values of significance for genetic variants for overweight MASLD along with genes near to the significant loci on chromosome 14 in the 93.8 to 94.8 Mb region. Genes containing significant loci are highlighted in bold and LLM-identified causal genes are colored red.

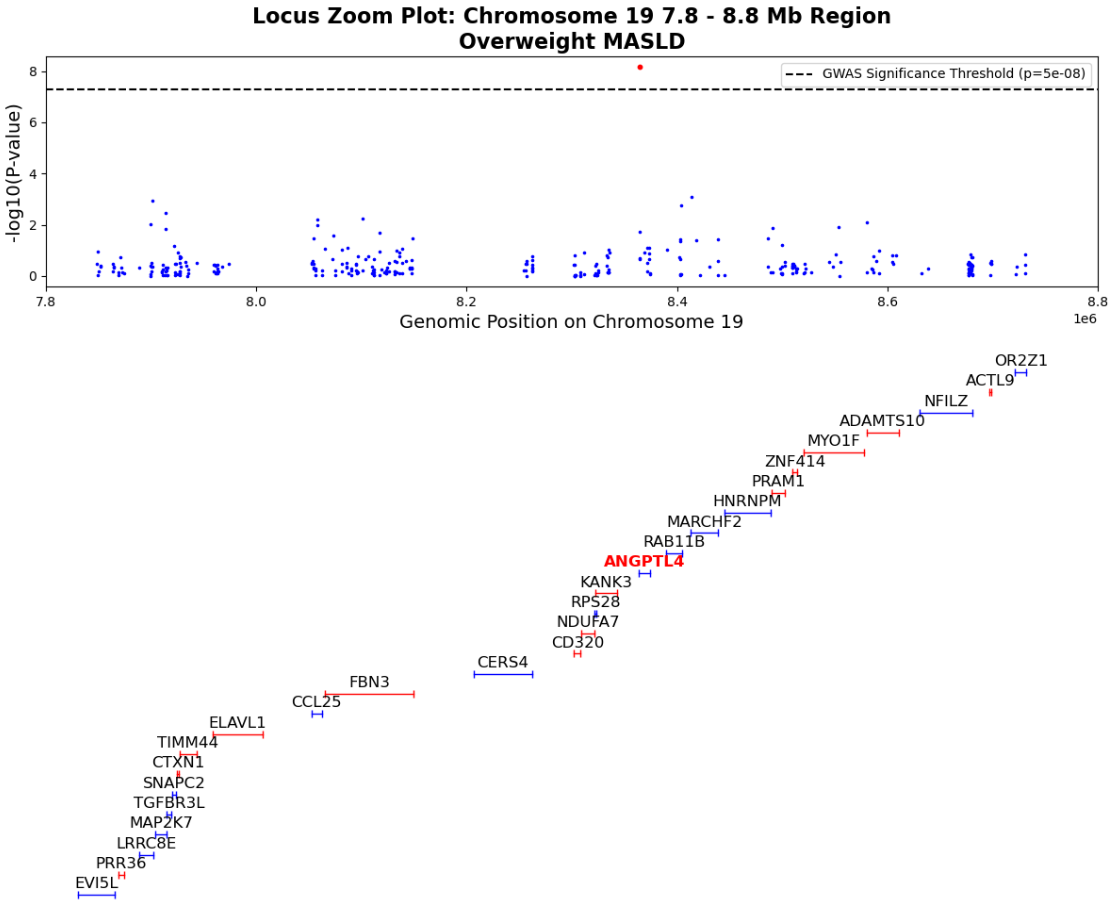


Figure 12: Locus zoom plot visualizing the p-values of significance for genetic variants for overweight MASLD along with genes near to the significant loci on chromosome 19 in the 7.8 to 8.8 Mb region. Genes containing significant loci are highlighted in bold and LLM-identified causal genes are colored red.

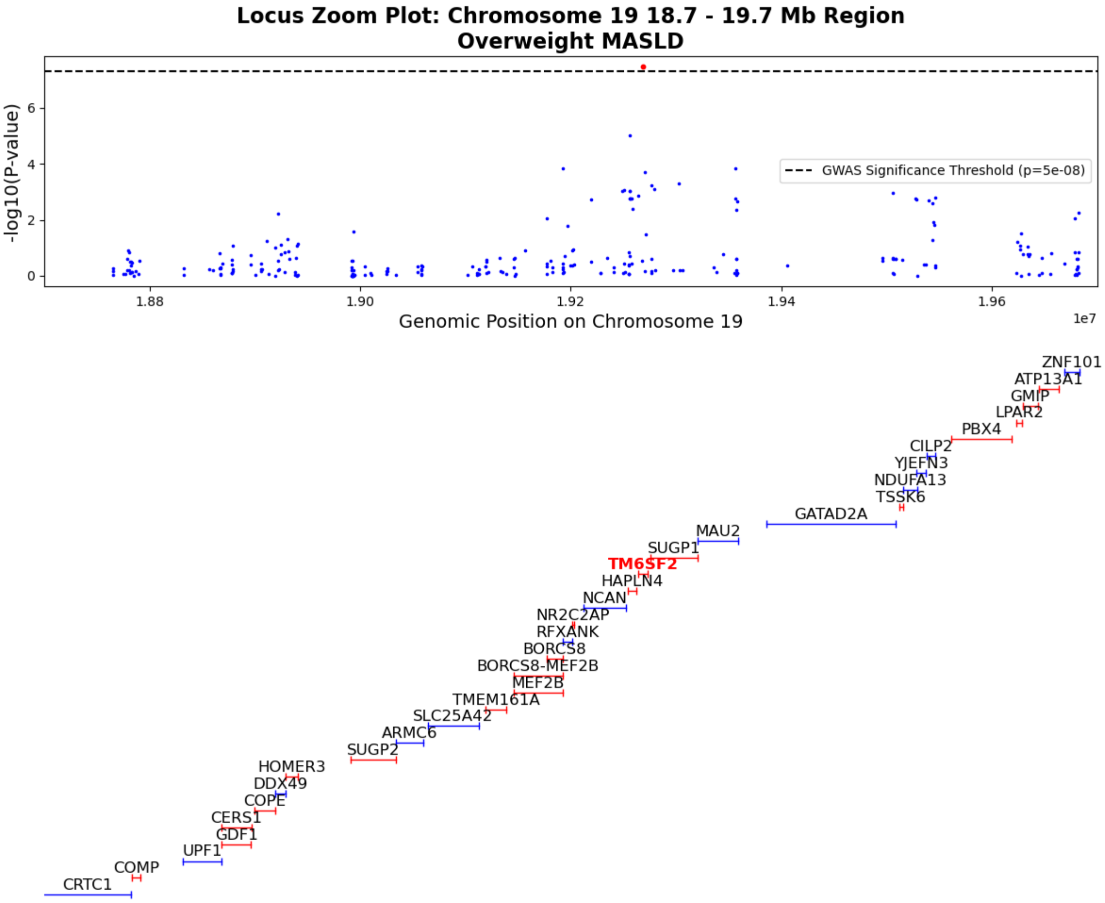


Figure 13: Locus zoom plot visualizing the p-values of significance for genetic variants for overweight MASLD along with genes near to the significant loci on chromosome 19 in the 18.7 to 19.7 Mb region. Genes containing significant loci are highlighted in bold and LLM-identified causal genes are colored red.

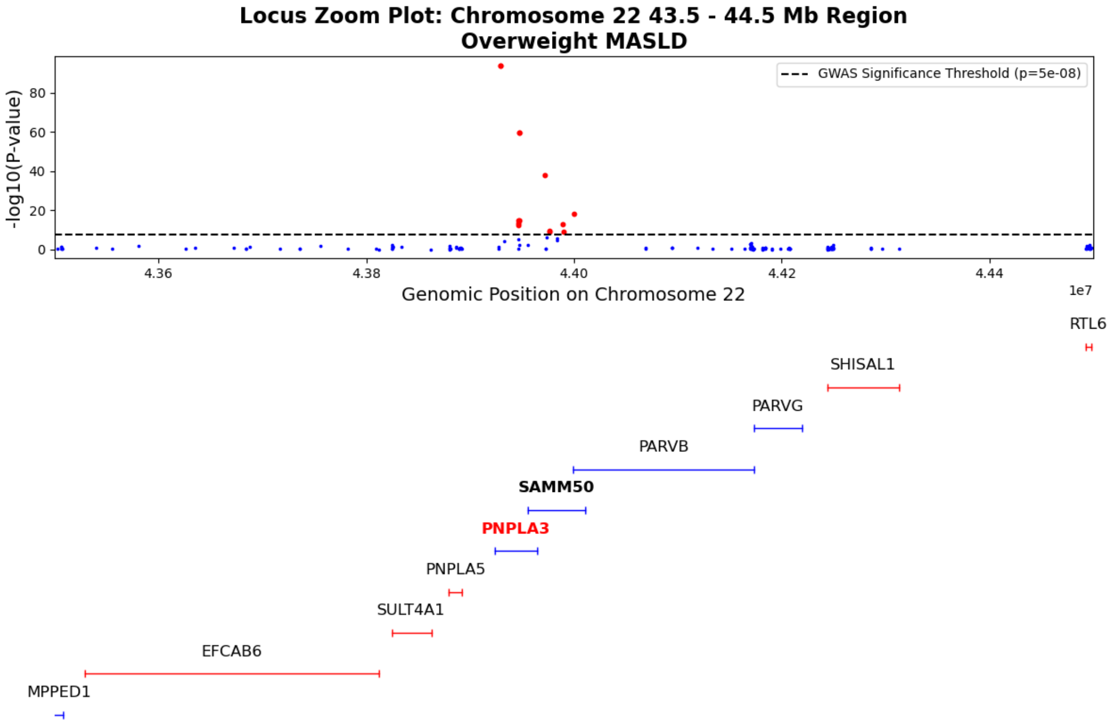


Figure 14: Locus zoom plot visualizing the p-values of significance for genetic variants for overweight MASLD along with genes near to the significant loci on chromosome 22 in the 43.5 to 44.5 Mb region. Genes containing significant loci are highlighted in bold and LLM-identified causal genes are colored red.

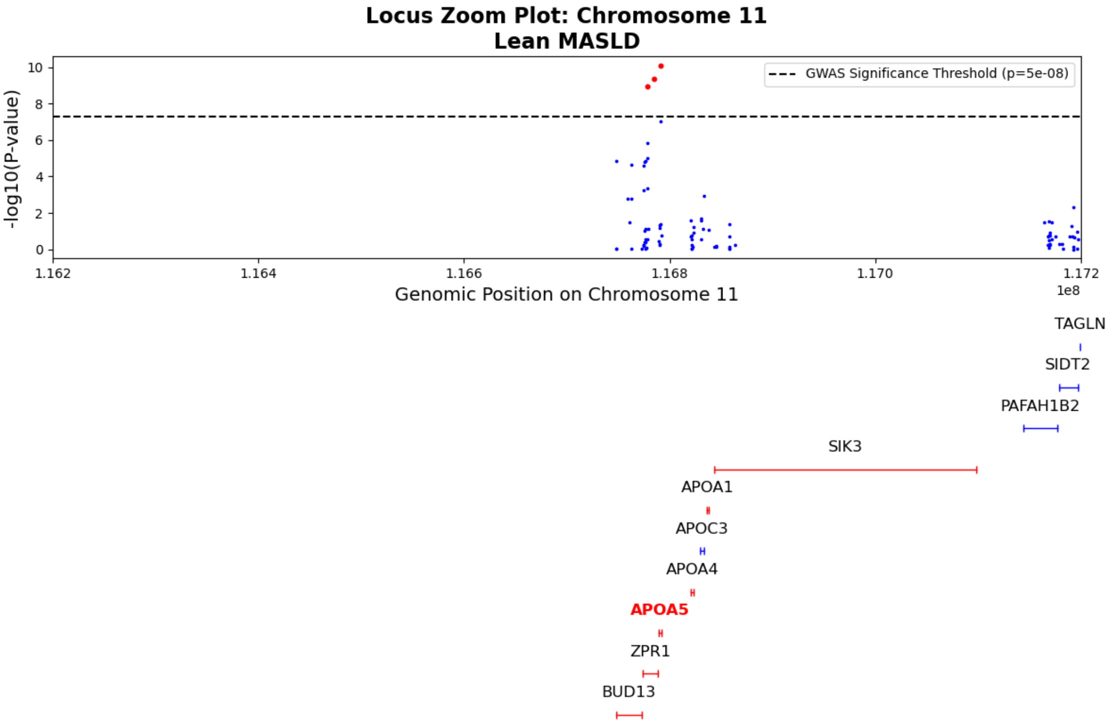


Figure 15: Locus zoom plot visualizing the p-values of significance for genetic variants for lean MASLD along with genes near to the significant loci on chromosome 11 in the 116.2 to 117.2 Mb region. Genes containing significant loci are highlighted in bold and LLM-identified causal genes are colored red.

| Feature Type | Feature |
|---|---|
| Laboratory measurements | Serum alanine aminotransferase<br>Serum aspartate aminotransferase<br>Serum glucose<br>Creatinine in urine<br>Cholesterol (HDL)<br>Triglyceride<br>Serum alkaline phosphatase<br>Serum albumin<br>Serum albumin-globulin ratio<br>Serum bilirubin<br>Cholesterol (LDL)<br>Serum creatinine |
| Vital signs | Systolic blood pressure<br>Diastolic blood pressure |
| Demographics | Race<br>Ethnicity<br>Sex assigned at birth |

Table 1: Features used to train the random forest model which predicts the I-MASLD scores.

| **Liver-Associated Condition** | **ICD9 Code(s)** | **ICD10 Code(s)** |
|---|---|---|
| MASLD | 571.5, 571.8, 571.9 | K76.0, K74.60, K74.69, K75.81, K75.89 |
| Alcoholic liver disease | 571.0-571.3, 303.00-303.03, 303.90-303.93 | K70.0-K70.9, F10.0, F10.120-10.129, F10.14, F10.150-10.159, F10.180-10.182, F10.188, F10.19, F10.20-10.21, F10.220-10.221, F10.229-F10.239, F10.24, F10.250-10.259, F10.26-10.27, F10.280-10.282, F10.288, F10.29 |
| Ascites | 785.59 | R18.8 |
| Spontaneous bacterial peritonitis | 567.23 | K65.2 |
| Cholangitis and Hemochromasis | 576.1, 275.01 | K83.0, K83.01, K83.09, E83.11, E83.110-83.111, E83.118-83.119 |
| Cirrhosis | 571, 571.5 | K70.3, K74.60, K74.69 |
| Diabetes | 250% | E08%, E09%, E10%, E11%, E13% |
| Encephalopathy | 572.2 | K72.91, G31.2, K76.82, K72.00, K70.4, K72.10, K71.1, K72.9, K70.41, K72.11, G62.1, K29.20, F10.988, K70.10, K70.30, K70.31, K70.0 |
| Esophageal Varices | 456.0, 456.1, 456.2, 456.20, 456.21 | I85.00, I85.01, I85.10, I85.11 |
| Hepatocellular Carcinoma | | C22.0, C22.9 |
| Hepatic Fibrosis | | K74.0, K74.00, K74.01, K74.02 |
| Hepatitis | 070.0-070.9, 072.71, 070.41, 070.44, 070.51, 070.54, 070.7-070.71, 571.4, 571.40-571.49, 573.1-573.2 | B17.10-B17.11, B18.2, Z22.52, A18.83, B00.81, B15.0, B15.9, B16.0-B19.9, B25.1, B58.1-B94.2, K70.10-K70.11, K71.2-K71.4, K71.50-K71.51, K71.6, K73.0-K73.9, K75.2-K75.4, O98.411-O98.413, O98.419, O98.42-O98.43, P35.3, Z22.50-Z22.52, Z22.59 |
| Liver Transplant | 996.8, 996.82, V42.7 | T86.40, T86.41, T86.42, T86.43, T86.49, Z94.4 |
| Metabolic and Nutritional Disease | | E88.81, E11.65, E11.9, E78.0, E78.1, E78.2, E78.5, E88.81 |

Table 2: Additional liver disease-associated ICD codes excluded from the controls cohort while training the I-MASLD model in the *All of Us* dataset.
